# Programmatic assessments implementation in a physiotherapy education curriculum – a study protocol for a randomized feasibility-controlled study

**DOI:** 10.64898/2026.02.28.26347309

**Authors:** Slavko Rogan, Narasimman Swaminathan, Jill Voegelin, Regina Cantieni, Patricia Wassmer, Simone Zingg, Eefje Luijckx

**Affiliations:** Division of Physiotherapy, School of Health Profession, Bern University of Applied of Sciences, Bern, Switzerland; School of Rehabilitation and Medical Sciences, University of Nizwa, Nizwa, Sultanate of Oman; Faculty of Movement Science and Exercise Rehabilitation, Vrije Universiteit Brussels, Brussels, Belgium

**Keywords:** Programmatic Assessment, Competence-Based Education, Physiotherapy Education, Learner-Centered Assessment, Feasibility Study

## Abstract

**Background:** Competence-Based Education (CBE) in physiotherapy aims to equip graduates with essential capabilities for safe and effective practice. Frameworks often include domains like clinical reasoning, communication, and professionalism. Despite its alignment with healthcare needs, CBE implementation in higher education remains inconsistent. Many educators still rely on behaviourist paradigms focused on passive learning and binary assessments, which inadequately reflect professional competence. Constructivist and progressive models offer more suitable alternatives yet are underutilized. Objective: This study explores the feasibility of integrating programmatic assessment (PA) to better support capability development and learner-centred education.

**Method:** This randomized controlled trial will be conducted a University of Applied Sciences across two campuses in Switzerland. Students from Cohort PHY25 enrolled in the BSc Physiotherapy program will be included. Students are assigned to PA in two formats, individual coaching (IG A) and group coaching (IG B), or to a sham PA without any coaching or reflective support (CG). Feasibility will be evaluated through session attendance, completion of all program components, and implementation fidelity. Secondary outcomes include staff readiness, wellbeing, workload, and learning gain.

**Discussion:** This study explores the feasibility and educational impact of implementing programmatic assessment in undergraduate physiotherapy education. If successful, PA may enhance competence development. Findings will inform curricular redesigns and support the shift toward learner-centred, capability-based assessment strategies in health professions education.

**Trial registration:** Registry of Efficacy and Effectiveness Studies under the number: #25261.2v1.

## Introduction

The world is undergoing profound transformations due to demographic shifts (1), environmental degradation, and economic instability. Population dynamics, such as size, composition, and spatial distribution are increasingly recognized as central to global environmental change and societal vulnerability (2). Overpopulation continues to exert pressure on ecosystems, contributing to deforestation, biodiversity loss, and climate instability (3, 4). Simultaneously, rapid population growth is linked to economic challenges including poverty, inequality, and resource scarcity, which further complicate sustainable development efforts (5). These global challenges are reshaping healthcare needs and placing increasing demands on health professionals to respond with agility, empathy, and competence. Higher education, particularly in health professions like physiotherapy, must adapt to this evolving landscape, not only by updating curricular content but also by rethinking pedagogical and assessment strategies (6).

Competence-based education (CBE) has emerged as a response to these demands, aiming to ensure that graduates possess the capabilities required for safe, effective, and context-sensitive practice (7-9). In physiotherapy, CBE frameworks emphasize domains such as clinical reasoning, communication, professionalism, and interprofessional collaboration, often operationalized through Entrustable Professional Activities (EPAs) (8). However, despite the theoretical alignment with modern healthcare needs, implementation in higher education remains inconsistent. A significant proportion of educators still rely on behaviourist paradigms, characterized by passive information transfer, imitation, and stimulus-response conditioning (10, 11). Assessments in such contexts are typically binary, using checklists or multiple-choice formats that fail to capture the complexity of professional competence.

Constructivist approaches (12, 13), which promote active learning and knowledge construction, are more widespread but still insufficiently integrated. Even less common are progressive models such as connectivism (14), andragogy (15), and heutagogy (16), which focus on the self-direction or self-determination of learners, self-organization, and context-related support. (17). These models align more closely with adult learning principles and the realities of clinical practice. In this context, assessment should not merely serve as a gatekeeping tool but as a driver of learning, a principle central to programmatic assessment (PA).

PA is a systems-based approach that collects multiple low-stakes and medium-stakes data points over time, integrating feedback/feedforward, mentoring, and reflection to inform high-stakes decisions (6, 18). It emphasizes proportionality (the richness of data should match the stakes of decisions) and meaningful triangulation (using diverse sources to assess competence holistically) (18). This approach supports both learning and robust decision-making, making it particularly suitable for capability development in health professions education.

The advantages of PA are well-documented. It fosters deeper learning, reduces exam-related stress, enhances feedback utilization, and supports early identification of struggling learners (6, 18). It also aligns with modern educational theories that emphasize learner agency and lifelong learning (19, 20). Institutions such as Maastricht University (Netherlands), Flinders University (Australia) (21), University of Fribourg (Switzerland) (22), and HAN University of Applied Sciences (Netherlands) (23) have successfully implemented programmatic assessment in medical and physiotherapy education.

The undergraduate physiotherapy program at a Swiss university of applied sciences is currently undergoing a pedagogical redesign to align with competency-based education (CBE) principles. This study protocol outlines the implementation of PA within this context, focusing on its feasibility and its impact on both students and staff.

The primary aim of this study is to evaluate the feasibility of implementing programmatic assessment in the undergraduate physiotherapy program. Feasibility will be assessed through 1. Session attendance, 2. program completion and 3. program fidelity (adherence to the intended assessment design)

Secondary outcomes will include staff readiness, student workload (self-reported and tracked), and learning gain (measured through pre-post self-assessments and portfolio analysis).

The research question is “to what extent is the implementation of programmatic assessment in the undergraduate physiotherapy program feasible, and how does it affect staff readiness, student’s workload, and learning outcomes?”

## Protocol

This manuscript follows the Reporting Guidelines SPIRIT 2010 statement for study protocols (24). Since we will be conducting a study at a higher education institution and not in a clinical setting, we have omitted the clinical points. Doi: 10.5281/zenodo.18621635 (25).

### Trial design

This study is planned as a randomized controlled trial at a Swiss university of applied sciences. The bachelor’s degree program in physical therapy is offered at two locations, one in Bern and the other in Basel. The study has been registered in the Registry of Efficacy and Effectiveness Studies under the number: 25261.1v1 from 20.09.2025 and re-registration due to host problem #25261.2v1 on 12.02.2026 (Link: https://sreereg.icpsr.umich.edu/sreereg/search/search).

### Trial setting

This study will be conducted at a School of Health Professions within a university of applied sciences in Switzerland, specifically within the Division of Physiotherapy. The study will involve students enrolled in the undergraduate physiotherapy cohort.

### Study status

Participant recruitment commenced on 15 September 2025 and was completed on 19 September 2025. Informed consent was obtained orally by the study nurse immediately following the informational session. Students were explicitly informed that participation was voluntary and that they could withdraw from the study at any time, including after initially providing consent. Following enrolment, students were randomized to the respective study groups using a predefined allocation procedure. The study is currently ongoing. Data collection is scheduled to be completed at the end of the Spring Semester 2026 (June), and the analysis and reporting of results are anticipated by the end of July 2026.

### Eligibility criteria

All students enrolled in the BSc Physiotherapy program and belonging to cohort PHY25 who are willing to participate in the study will be eligible for inclusion. Students from other cohorts, as well as those who do not provide informed consent, will be excluded from participation.

### Intervention and Comparator

The study will be conducted at two locations offering bachelor’s degree programs in physical therapy. Students will be randomly assigned to one of three groups: the PA intervention with individual coaching (IG A), the PA intervention with group coaching (IG B), or the control group receiving a sham PA. The intervention will take place during the first semester of the 2025/26 academic year and will include three structured data collection points:

- Data Point 1 (calendar weeks 44–46): Progress test
- Data Point 2 (weeks 49–50): Practical skills test in triads (patient, therapist, observer) with peer feedback
- Data Point 3 (weeks 03–04): Second progress test

After each data point, students in the intervention groups (IG A and IG B) receive coaching— individually in IG A or in groups in IG B—and complete a written reflection including learning goals. The final module examination in week 5 is an Objective Structured Clinical Examination (OSCE). The control group (CG) completes the same data points but does not receive coaching or any additional support. A visual timeline of the study is provided in Figure 1.

**figures 1.** Study flow.

### Outcomes

The feasibility of the PA implementation will be evaluated based on three primary indicators: session attendance, program completion, and program fidelity.

In addition, secondary outcomes will be assessed, including staff readiness, student workload, wellbeing, and learning gain. Table 1 present the primary and secondary outcomes.

**Table 1.** Study target.

| Target Outcome |  | What to measure | Assessment outcome |
| --- | --- | --- | --- |
| Primary Outcome |  |  |  |
|  | Criteria of success |  |  |
|  | Data point session attendance | How many students attended each data point session of the programme? | Number of students |
|  | Programme completion | How many students participate at all 3 data points | Number of students in % (80) |
|  | Programme fidelity | How many data points were completed as planned? For data point 2, observers record in observation templates whether a session was completed as planned or not. The observer is instructed to note any circumstances that prevented the session from being completed as planned. | Adherence % (target 80%)<br>Attrition % (target 0%) |
| Secondary outcome |  |  |  |
|  | Level 1 |  |  |
|  | Readiness | Staff members | Questionnaire Survey<br>Evasys tool<br>Group interview |
|  | Workload | Students working hours per week: Preparation, lecture, seminars, self-study time, follow-up, | hours |
|  | Level 2 |  |  |
|  | Learning gain | Objective structure clinical examination (OSCE) | Grades, reflexion |
|  | Well-being | Mental Health (German Warwick-Edinburgh Mental Well-being Scale) | Likert scale |

### Sequence generation

Random allocation was used to assign students to the three study groups. All eligible students in cohort PHY25 were randomized into one of the following groups: (1) PA with individual coaching, (2) PA with group coaching, or (3) sham-PA control. Randomization was conducted at the individual student level using a computer-generated random sequence. No stratification or blocking procedures were applied.

### Allocation concealment mechanism

The random allocation sequence was generated by an independent study administrator who was not involved in teaching or assessment. Allocation was implemented using a central, password-protected digital system. The group assignments were released only after students had been irrevocably enrolled in the study, ensuring full allocation concealment until the moment of assignment.

### Blinding

Due to the nature of educational intervention, blinding of educators and students is not feasible. However, to minimize bias, both the outcome assessor and the data analyst will remain blinded to group allocation throughout the study.

### Data collection methods

Data collection for this study will occur between October 2025 and May 2026. Data point session attendance will be evaluated using institutional records, such as Moodle (Moodle Pty Ltd, Perth, Australia) or UCAN (Umbrella Consortium for Assessment Networks, Heidelberg, Germany) tool login data, and attendance lists. Program completion and program fidelity will be evaluated using Moodle and Evasys (Electric Paper Evaluationssysteme GmbH, Lüneburg, Germany) attendance logs, as well as teacher attendance logs. Program fidelity will be evaluated using Moodle 4.2.x and Evasys (V9.1 (2469), 2023) attendance protocols as well as attendance logs from instructors. Staff member readiness will be evaluated using validated questionnaires and semi-structured interviews. Student workload will be assessed through weekly self-report logs and the NASA Task Load Index. To ensure data quality, assessors will receive standardized training, and duplicate measurements will be taken where appropriate. All instruments used are supported by literature, and data collection forms will be made available via the institutional repository or upon request.

### Data management

Data will be securely stored and coded. Trained assessors will enter data using range checks and double entry for accuracy. Access is restricted, and procedures follow institutional data protection standards.

### Statistics

The primary independent variable in this study is the type of assessment model: PA with coaching and reflection versus sham PA without coaching or reflection. Dependent variables include feasibility indicators (session attendance, program completion, program fidelity) and secondary outcomes (staff readiness, student workload, and learning gain).

Descriptive statistics will be used to summarize baseline characteristics and feasibility outcomes. Group comparisons will be conducted using appropriate inferential tests (e.g. non-parametric equivalents), depending on data distribution. To account for potential confounding, multivariable regression models will be applied, adjusting for covariates such as baseline academic performance, age, gender, prior clinical experience, motivation, teaching staff experience, and site-specific factors.

Missing data will be handled using multiple imputations where appropriate. Subgroup analyses will explore differences between individual and group coaching formats. Sensitivity analyses will assess the robustness of findings across different model specifications.

## Discussion

This implementation study explores the feasibility and educational impact of implementing a programmatic assessment model in undergraduate physiotherapy education. The results will provide insight into how structured, longitudinal assessment combined with coaching and reflection can support competence development in a real-world academic setting. By comparing outcomes between the intervention and control groups, the study aims to identify strengths and challenges in applying PA within a physiotherapy curriculum.

Feasibility indicators such as session attendance, program completion, and fidelity will help determine whether the PA model is practical and sustainable. Secondary outcomes, including staff readiness, workload, and learning gain, will offer a broader understanding of how PA affects the learning environment and student experience. The inclusion of covariates such as prior academic performance, motivation, and site-specific factors will allow for a nuanced interpretation of the results.

If successful, this study could inform broader curricular redesigns and support the integration of competence-based, learner-centered assessment strategies in health professions education. It may also contribute to the growing body of literature advocating for assessment as a driver of learning rather than a mere evaluative tool.

## Dissemination

Findings from this study will be disseminated through multiple channels. An internal summary report will be shared with faculty members and curriculum committees at the institution. Results will be presented at national and international conferences in health professions education, and a full manuscript will be submitted to a peer-reviewed journal. In addition, key insights will be communicated to physiotherapy educators across Switzerland and Europe to support evidence-informed teaching and assessment practices.

## Data Availability

No datasets were generated or analysed during the current study.

## Ethical considerations

All participants will give their informed consent in accordance with the standards of ethics. This study falls under the Human Research Act, Category A, low risk. This study protocol has already been approved by the cantonal ethics committee of Bern (Req-2025-00706).

## Data availability

Rogan, S. R. (2026). SPIRIT Checklist for Programmatic assessments implementation in a physiotherapy education curriculum – a study protocol for a randomized feasibility-controlled study (Version 1). Zenodo: Licence; Creative Commons Attribution 4.0 International; . https://doi.org/10.5281/zenodo.18621635

## Competing interests

Authors declare no competing interests

## Grant information

None

## Acknowledgments

We have no acknowledgments to make.

## Notes

### Competing Interest Statement

The authors have declared no competing interest.

### Funding Statement

The author(s) received no specific funding for this work.

